# Analysis clinical features of COVID-19 infection in secondary epidemic area and report potential biomarkers in evaluation

**DOI:** 10.1101/2020.03.10.20033613

**Authors:** Weiping Ji, Gautam Bishnu, Zhenzhai Cai, Xian Shen

## Abstract

**Objective:** Based on the clinical characteristics of infected patients with novel coronavirus in secondary epidemic areas, we aimed to identify potential biomarkers for the evaluation of novel coronavirus-infected patients, guide the diagnosis and treatment of this disease in secondary epidemic areas and provide a reference for the clinical prevention and control of this epidemic situation.

**Methods:** The clinical data of 33 patients with respiratory symptoms caused by the novel coronavirus in Wenzhou city from January 15 to February 12, 2020, were thoroughly reviewed. At the onset of the disease, we found that the primary symptoms were fever, cough, fatigue, chest tightness, chest pain and specific blood test results. According to the patients’ histories, the patients were divided into two groups: those who spent time in the main epidemic area and those who did not spend time in the main epidemic area. The differences in the clinical manifestations between these two groups were analyzed.

**Results:** The main clinical symptoms of patients infected with novel coronavirus in the secondary epidemic area were respiratory tract ailments and systemic symptoms. After grouping patients based on the presence or absence of residency in or travel history to the main epidemic area, there was no significant difference between the baseline data of these two groups, and there were no significant differences in symptoms and signs between the two groups (P>0.05). Some patients had abnormally increased serum amyloid protein A (SAA). There were statistically significant differences in the leukocyte count/C-reactive protein, monocyte ratio/C-reactive protein, neutrophil count/C-reactive protein, monocyte count/C-reactive protein and hemoglobin/C-reactive protein values between the two groups (P < 0.05).

**Conclusion:** Respiratory tract ailments and systemic symptoms were the primary symptoms of novel coronavirus infection in the secondary epidemic area; these symptoms are not typical. The abnormal increase in serum amyloid protein (SAA) may be used as an auxiliary index for diagnosis and treatment. CRP changes before other blood parameters and thus may be an effective evaluation index for patients with COVID-19 infection.

---

A new viral pneumonia disease outbreak in Wuhan city at the beginning of 2020 was confirmed to be caused by the “COVID-19” novel coronavirus infection [1]. COVID-19 has not been previously found in the human body, and no previous case of COVID-19 pneumonia has been detected. The primary clinical symptoms of those infected with coronavirus are chills, severe acute respiratory syndrome (SARS) and Middle East respiratory syndrome (MERS) [2]. According to a report from Wuhan city, the main epidemic area, people infected with COVID-19 often have symptoms of fever, cough, shortness of breath, dyspnea, and severe infection, which can lead to pneumonia or even death [3]. The epidemic has spread from the main epidemic areas, Hubei Province and Wuhan city, to other provinces and cities, where new successive cases of COVID-19 infection have been reported. In some cities, such as Wenzhou city of Zhejiang Province, the epidemic situation is serious. Therefore, these locations outside the main epidemic area have been designated secondary epidemic areas**. In this study, the first cluster of cases of COVID-19 infection in Wenzhou city, Zhejiang Province, which is the leading secondary epidemic area, is reported.

**Note: Secondary epidemic areas refer to new areas that are experiencing epidemics caused by the arrival of the infected patients from main epidemic areas and in which the disease is spreading, similar to the main epidemic areas of Hubei Province and Wuhan city.

## 1. Data and Methods

### 1.1 Clinical Data

The clinical data of patients who were diagnosed with novel coronavirus COVID-19 positive by nucleic acid test from sputum, throat swab, lower respiratory secretion and other samples were collected from January 15 to February 1, 2020. All patients gave signed, informed consent for their dates to be used for scientific research. Ethical approval for the study was obtained from the Second Affiliated Hospital of Wenzhou Medical University (Zhejiang, China).

### 1.2 Inclusion criteria

Patients aged 18 years or older who met the NCP diagnostic criteria were included [4].

### 1.3 Exclusion criteria

⍰ Patients with non-COVID-19 infectious respiratory diseases; and ⍰ patients with suspected COVID-19 but multiple negative sputum, throat swab, lower respiratory secretion and other samples were excluded.

### 1.4 Grouping method

According to the previous histories of the patients who met the COVID-19 infection diagnostic criteria, the cases that occurred after returning to Wenzhou from Wuhan were assigned to the direct contact (DC) group. Those with no history of residence in Wuhan or the main epidemic area before the onset of the disease were assigned to the indirect contact (IDC) group.

### 1.5 Clinical and test indicators

Symptoms such as fever, cough, fatigue and soreness at the time of examination and the results of tests were compared between the two groups.

### 1.6 Statistical method

SAS 9.4 statistical analysis software was used. The t-test was used for comparisons between groups for continuous variables. The comparison between groups for categorical variables was performed using Fisher’s exact probability method. Means and standard deviations are used to describe centralized discrete trends. The t-test, rank-sum test, and Fisher’s exact probability method were used for comparisons between groups. P≤0.05 indicates that the difference was statistically significant.

## 2 Results

### 2.1

According to the descriptions of basic symptoms and signs of the total study population, the data of 33 conformed COVID-19 cases, including 17 males (51.5%) and 16 females (48.5%), were analyzed. The DC group contained 15 patients, nine males (60%) and six females (40%), with an average age of 44.67 ± 12.62 years. In the IDC group, there were 18 patients, eight males (44.4%) and ten females (55.6%), with an average age of 47.11 ± 10.09 years; In the DC group, 2 patients (20%) had hypertension, 3 patients (30%) had hyperglycemia, and 1 patient (10%) had hyperlipemia. In the IDC group, 4 (22.2.%) patients had hypertension, 0 patients (0%) had hyperglycemia, and 0 patients (0%) had hyperlipemia. The DC group, the average body temperature was 37.97 ± 0.36, and 15 cases had fever (100%), 10 cases had sore throat (33.3%), 10 cases had cough (33.3%), 9 cases had cough (60%), 7 cases has nasal congestion (46.7%), 8 cases (53.3%) had fatigue, and 6 cases (40%) had muscle soreness. In the IDC group, the average body temperature was 38.14 ± 0.67°, and there was fever in 17 cases (94.4%), sore throat in 3 cases (16.7%), cough in 14 cases (77.8%), productive cough in 8 cases (44.4%), nasal congestion in 2 cases (11.1%), fatigue in 8 cases (44.4%), and muscle soreness in 5 cases (27.8%). There were no significant differences between the groups. See Tables 1 for other data on nausea, vomiting, respiratory distress and chest pain.

### 2.2

The comparison of blood test results and C-reactive protein (CRP) test results in the two groups showed that the average white blood cell count in the DC group was 4.90 (3.55-6.38) × 10^9^ /L, the neutrophil count was 3.02 ± 1.38 × 10^9^ /L, the lymphocyte count was 1.24 ± 0.45 × 10^9^ /L, the monocyte count was 0.44 (0.30-0.78) × 10^9^ /L, the eosinophil count was 0.01 (0.00-0.02) × 10^9^ /L, the eosinophil basic granulocyte count was 0.01 (0.01-0.02) × 10^9^ /L, hemoglobin (Hb) was 149.50 ± 15.15 g/L, the red blood cell count was 4.83 ± 0.53 × 10^9^ /L, the platelet (PLT) count was 172.14 + 36.42 × 10^9^ /L, and C-reactive protein (CRP) was 5.68 (2.80-13.00) mg/L. In the IDC group, the average white blood cell count was 4.45 (3.77-6.15) × 10^9^ /L, the neutrophil count was 2.98 ± 1.20 ×10^9^ /L, the lymphocyte count was 1.25 ± 0.30 × 10^9^ /L, the monocyte count was 0.32 (0.24-0.48) × 10^9^ /L, the eosinophil count was 0.01 (0.00-0.02) × 10^9^ /L, the eosinophil basic granulocyte count was 0.01 (0.01-0.02) × 10^9^ /L, hemoglobin (Hb) was 143.33 ± 15.96 g/L, the red blood cell count was 4.93 ± 0.44 × 10^9^ /L, the platelet (PLT) count was 187.62 ± 39.55 × 10^9^ /L, and C-reactive protein (CRP) was 11.89 (9.74-23.36) mg/L; neutrophil ratios and other data are shown in Table 2.

### 2.3

Serum amyloid protein A (SAA) test: Among the 33 patients enrolled in the study, 5 patients in the DC group were tested for serum amyloid protein A (SAA). It was found that SAA in 4 patients was increased significantly, with an average value of 61.51 mg/L (normal value ≤ 10) mg/L). The increase in this protein was more obvious than the increase in CRP, but due to the limited data, a comparative statistical analysis was not performed. In the future, we will focus on the SAA of NCP patients. See Table 3 for the detailed data.

### 2.4

Comparison of the blood cell count to C-reactive protein (CRP) ratio: In the DC group, the leukocyte count/C-reactive protein, monocyte ratio/C-reactive protein, neutrophil count/C-reactive protein, monocyte count/C-reactive protein, and hemoglobin/C-reactive protein values were 0.55 (0.42-1.89), 0.02 (0.01-0.05), 0.39 (0.29-1.02), 0.07 (0.05-0.18), and 23.20 (11.54-56.79), respectively. In the IDC group, the leukocyte count/C-reactive protein, monocyte ratio/C-reactive protein, neutrophil count/C-reactive protein, monocyte count/C-reactive protein, and hemoglobin/C-reactive protein values were 0.31 (0.19-0.86), 0.01 (0.00-0.01), 0.18 (0.10-0.40), 0.02 (0.01-0.05), and 11.51 (4.75-16.67), respectively. The differences between the blood cell counts and C-reactive protein ratios between the two groups were statistically significant (all P < 0.05). See Table 4 for detailed data.

## 3 Discussion

According to the reports of scholars in the main epidemic area, the primary clinical symptoms of COVID-19-infected patients are fever, dry cough, fatigue, and the gradual development of respiratory problems, with fever being the most typical. Only few patients will present with obvious upper respiratory symptoms (e.g., runny nose, sneezing and sore throat), and diarrhea, headache and hemoptysis are rare [5]. Patients with mild infection may show only slight fatigue and no fever [6]. Critically ill patients may develop symptoms such as acute respiratory distress syndrome, septic shock, metabolic acidosis, and coagulation dysfunction [7] in addition to symptoms such as shortness of breath, audible wet sounds in both lungs, and weak breath sounds. Dullness on percussion and tactile tremor may be increased or decreased [8]. In our study, we found that the clinical symptoms in COVID-19-infected patients in the secondary epidemic area were similar to those in patients in the primary epidemic area, but there were some inconsistencies. In the secondary epidemic area, the main symptoms were fever, mainly moderate- to low-grade fever. Cough was not usually dry and was often accompanied by white phlegm. Fatigue and muscle aches were not seen in most patients. The symptoms of upper respiratory tract infection (such as sneezing, runny nose, sore throat and itching) were not uncommon. The symptoms of chest distress and chest pain were less common in the secondary epidemic area than in the main epidemic area; this may be related to the rapid diagnosis in the secondary epidemic area and early treatment and intervention. Moreover, critical patients were rare, which may be related to the younger age of patients and the presence fewer complications in the secondary epidemic area than in the main epidemic area.

According to a report from the main epidemic area, COVID-19-infected patients had limited lung lesions at the early stage of onset. CT showed multiple lesions and rarely single lesions in the lungs. CT findings appeared as localized small patches, sub-segmented or large ground glass shadows. Local leaflets were widely spaced and were with or without thickening. Progressive CT showed increased lesions, with some expanded lesions, and multiple lung lobes were involved; the density of the lesions increased, and irregular, wedge-shaped or fan-shaped consolidations of varying sizes and degrees within the lesions appeared. Mutated shadows or strip shadows coexisted. Diffuse lung lesions can be seen on CT in severe patients, with consolidation as the primary cause, in addition to ground glass shadows and multiple fiber strand shadows; in a few severe cases, “white lung” manifestations appeared when most of the lungs were affected, and an air bronchus sign was present [9-12]. The results of imaging examinations of 33 patients in the secondary epidemic area collected in this study were similar to those reported in the main epidemic area, so no further explanation is given in the article.

In patients with COVID-19 infection diagnosed in the main epidemic area, the laboratory examination results at the early stage of the disease showed that the total number of white blood cells decreased or remained normal; the lymphocyte count decreased; the monocyte count increased or remained normal; the liver enzyme, muscle enzyme and myoglobin levels increased in some patients; the C-reactive protein and ESR increased in most patients; procalcitonin remained normal; D-dimer increased in severe patients; lymphocytes progressively decreased; and coagulation function decreased. Inflammatory cytokines, such as interleukin-2, tumor necrosis factor-α (TNF-α), IL-6, and interferon-γ (IFN-γ), remained normal or slightly increased. The level of cytokines in patients with organ failure was significantly increased. In addition, coronavirus nucleic acids were detected in throat swabs, sputum samples, lower respiratory secretions and blood samples [6,7]. Among the infected patients, the viral load detected in the secretion of the lower respiratory tract was higher than that detected in the upper respiratory tract [9,13]. The laboratory examination results of the 33 patients showed that the total number of leukocytes in patients with COVID-19 infection in the secondary epidemic area was slightly reduced, normal or slightly increased, and the proportion of lymphocytes in some patients was significantly reduced. Rarely, lymphocytes or neutrophils were increased, and monocytes and eosinophilic acid-base granulocytes were not changed significantly. CRP did not change significantly in all patients with COVID-19 infection, and some patients maintained a normal CRP level, but it was found that serum amyloid A (SAA) increased significantly in most patients that were tested; however, due to the limited amount of data, a statistical analysis was not performed. SAA is an acute-phase protein. SAA significantly increases in viral infection, whereas CRP may not increase or only slightly increase in viral infection without bacterial infection. Moreover, SAA increases in both viral and bacterial infections, and the increase reflects the severity of the infection. At present, it is generally agreed that SAA and CRP should be combined to judge inflammatory activity [14]. Considering that COVID-19 in the secondary epidemic area may occur after the primary epidemic in the main epidemic area, and its toxicity and pathogenicity may be weakened, SAA detection in the secondary epidemic area should be considered to evaluate the occurrence and development of COVID-19 infection. We will focus on the detection of SAA in COVID-19-infected people in follow-up studies.

The significant differences in the ratios between various blood parameters and CRP between the two groups are interesting findings in this study. It has been reported that CRP may not increase or only slightly increase in viral infection without bacterial infection, and the increase indirectly reflects the severity of infection. At present, it is generally accepted that CRP or combined CRP can be used to judge the inflammatory activity [15]. In this study, we found that the ratio between various blood parameters and CRP was significantly increased in patients who had indirect contact with the disease in the epidemic area, suggesting that the change in the reactivity of CRP in COVID-19 infection is greater than the response of various blood parameters. In addition, considering that COVID-19 in the secondary epidemic area may occur after the primary epidemic in the main epidemic area, its toxicity and pathogenicity may be weakened. CRP can be used as a predictive factor prior to changes in leukocytes, lymphocytes, neutrophils and other inflammatory-related blood parameters to comprehensively evaluate the occurrence and development of COVID-19 infection.

## Authors’ contributions

Manuscript was designed and completed by Weiping Ji, Gautam Bishnu completed data and language work, Zhenzhai Cai completed statistics and verification, Xian Shen completed the review and revision of the paper, and guided the whole process.

## Acknowledgements

The authors declared no competing interests exist.

## References

[1] Zhu N, Zhang D, Wang W, et al. A novel coronavirus from patients with pneumonia in China,N Engl J Med. 2020 Jan 24. doi: 10.1056/NEJMoa2001017. [Epub ahead of print]

[2] Killerby ME, Biggs HM, Midgley CM, Gerber SI, Watson JT.Middle East Respiratory Syndrome Coronavirus Transmission.Emerg Infect Dis. 2020;26(2):191-198.

[3] Liu X, Zhang J, Wang BN, et al. Early Transmission Dynamics in Wuhan, China,of Novel Coronavirus–Infected Pneumonia.Engl J Med. 2020 Jan 29. doi: 10.1056/NEJMoa2001316. [Epub ahead of print]

[4] Victor M Corman, Olfert Landt, Marco Kaiser, et al.Detection of 2019 novel coronavirus (COVID-19) byreal-time RT-PCR.Euro Surveill. 2020 Jan;25(3). doi:10.2807/1560-7917.ES.2020.25.3.2000045.

[5] Chaolin Huang, Yeming Wang, Xingwang Li, et al. Clinical features of patients infected with 2019 novelcoronavirus in Wuhan, China.Lancet. 2020 Jan 24. pii: S0140-6736(20)30183-5. doi: 10.1016/S0140-6736(20)30183-5. [Epub ahead of print]

[6] Wang D, Hu B, Hu C, et al. Clinical Characteristics of 138 Hospitalized Patients With 2019 Novel Coronavirus-Infected Pneumonia in Wuhan, China. JAMA. 2020 Feb 7. doi: 10.1001/jama.2020.1585. [Epub ahead of print]

[7] Chen N S, Zhou M, Dong X, et al. Epidemiological and clinical characteristics of 99 cases of 2019 novel coronavirus pneumonia in Wuhan,China:a descriptive study[J]. Lancet,2020,doi: 10.1016/S0140-6736(20)30211

[8] Jin Yinghui, Cai Lin, Wang xinghuan, et al. A guidelines for rapid recommendations for the diagnosis and treatment of Novel coronavirus (COVID-19) infection (Standard Version) [J/OL].PLA medical journal: 1–20

[9] Chan JF, Yuan S, Kok KH. et al..A familial cluster of pneumonia associated with the 2019 novel coronavirus indicating person-to-person transmission: a study of a family cluster. Lancet. 2020 Jan 24. pii: S0140-6736(20)30154–9. doi: 10.1016/S0140-6736(20)30154-9. [Epub ahead of print]

[10] Shi He Shui, Han Xiaoyu, Fan Yanqing. et al. Clinical Features and Imaging Features of Pneumonia Infected by New Coronavirus (COVID-19). Journal of Clinical Radiology, 2020.

[11] Guan Hanxiong, Xiong Ying, Shen Nanxi. et al. Preliminary study on clinical imaging characteristics of Wuhannew 2019 coronavirus (COVID-19) pneumonia .radiology practice, 2020

[12] Li B, Si HR, Zhu Y, et al. Discovery of Bat Coronaviruses through Surveillance and Probe Capture-Based Next-Generation Sequencing. mSphere. 2020 Jan 29;5(1). pii: e00807-19. doi: 10.1128/mSphere.00807-19.

[13] Zhang N, Wang L, Deng X, et al.Recent advances in the detection of respiratory virus infection in humans. J Med Virol. 2020 Jan 15. doi: 10.1002/jmv.25674. [Epub ahead of print]

[14] Yip TT, Chan JW, Cho WC et al. Protein chip array profiling analysis in patients with severe acute respiratory syndrome identified serum amyloid a protein as a biomarker potentially useful in monitoring the extent of pneumonia.Clin Chem. 2005;51(1):47–55.

[15] dos Anjos BL, Grotto HZ.Evaluation of C-reactive protein and serum amyloid A in the detection of inflammatory and infectious diseases in children. Clin Chem Lab Med, 2010; 48:493–499.

